# A point-of-care lateral flow assay for neutralising antibodies against SARS-CoV-2

**DOI:** 10.1101/2021.04.12.21255368

**Authors:** Thomas S. Fulford, Huy Van, Nicholas A. Gherardin, Shuning Zheng, Marcin Ciula, Heidi E. Drummer, Samuel Redmond, Hyon-Xhi Tan, Rob J. Center, Fan Li, Samantha L. Grimley, Bruce D. Wines, Thi H.O. Nguyen, Francesca L. Mordant, Louise C. Rowntree, Allen C. Cheng, Denise L. Doolan, Katherine Bond, P. Mark Hogarth, Zoe McQuilten, Kanta Subbarao, Katherine Kedzierska, Jennifer A. Juno, Adam K. Wheatley, Stephen J. Kent, Deborah A. Williamson, Damian F.J. Purcell, David A. Anderson, Dale I. Godfrey

## Abstract

As vaccines against SARS-CoV-2 are now being rolled out, a better understanding of immunity to the virus; whether through infection, or passive or active immunisation, and the durability of this protection is required. This will benefit from the ability to measure SARS-CoV-2 immunity, ideally with rapid turnaround and without the need for laboratory-based testing. Current rapid point-of-care (POC) tests measure antibodies (Ab) against the SARS-CoV-2 virus, however, these tests provide no information on whether the antibodies can neutralise virus infectivity and are potentially protective, especially against newly emerging variants of the virus. Neutralising Antibodies (NAb) are emerging as a strong correlate of protection, but most current NAb assays require many hours or days, samples of venous blood, and access to laboratory facilities, which is especially problematic in resource-limited settings. We have developed a lateral flow POC test that can measure levels of RBD-ACE2 neutralising antibodies from whole blood, with a result that can be determined by eye (semi-quantitative) or on a small instrument (quantitative), and results show high correlation with microneutralisation assays. This assay also provides a measure of total anti-RBD antibody, thereby providing evidence of exposure to SARS-CoV-2 or immunisation, regardless of whether NAb are present in the sample. By testing samples from immunised macaques, we demonstrate that this test is equally applicable for use with animal samples, and we show that this assay is readily adaptable to test for immunity to newly emerging SARS-CoV-2 variants. Lastly, using a cohort of vaccinated humans, we demonstrate that our whole-blood test correlates closely with microneutralisation assay data (R^2^ =0.75, p<0.0001), and that fingerprick whole blood samples are sufficient for this test. Accordingly, the COVID-19 NAb-test™ device described here can provide a rapid readout of immunity to SARS-CoV-2 at the point of care.

## Introduction

The COVID-19 pandemic is having a major impact on human activity and well-being. The causative agent, SARS-CoV-2, has infected over 130 million people and caused over 2.8 million deaths, with many more left with potentially lasting damage to lungs and other organs^1, 2^. The first vaccines are now being rolled out in many countries after a highly accelerated development process, and rapid completion of clinical efficacy trials in the setting of high disease incidence, with results from phase III trials and real world studies suggesting efficacy rates of up to 95%^3^. Whether these vaccines can prevent infection and transmission is less clear, but their ability to prevent disease is already a major advance.

Despite this rapid progress, it is unlikely that COVID-19 will be eradicated in the near future and at best, the hope is that COVID-19 infections can be brought to a level of control where life can return to a “post-COVID” normality. A key part of this solution will be widespread vaccination, together, in some circumstances, with testing to determine who is immune, whether through prior infection or vaccination, and how durable immunity is, both on a population level and on an individual basis.

Antibody responses to viral infections and/or vaccines are typically determined using serological tests such as ELISAs or point-of-care (POC) lateral flow assays. These assays measure virus-specific antibody responses, that include immunoglobulin subclasses such as IgG, IgM or IgA. Such assays are widely available for the detection of anti-SARS-CoV-2 antibodies, with variable levels of specificity and sensitivity^4^. For some viruses such as hepatitis A and hepatitis B, where the protective epitopes are highly immunodominant and demonstrate minimal variation, total antibody tests provide a satisfactory correlate of antibody-based immunity. However, for SARS-CoV-2 and many other viruses, the combination of adaptive mutations to enhance receptor binding affinity, immune selection and at least moderate error rates during replication can lead to the rapid selection of variants that may escape the antibody response (akin to “antigenic drift” in influenza viruses). This is exemplified by the recent emergence of viral variants of concern (VOC) such as the B.1.351 lineage first detected in South Africa. Recent evidence suggests that some current vaccines may show signs of reduced efficacy against such variants because the variants are not efficiently neutralised by antibodies induced by the vaccine^5^. This will require not only the development of modified vaccines, but also the development of serological tests that can detect and quantify the dominant neutralizing antibody responses to newly emerging strains.

Neutralising antibodies are emerging as a strong correlate of protection^6, 7^ but are only a subset of the total polyclonal response. The dominant target of NAb in SARS-CoV-2 is the Receptor Binding Domain (RBD), located at the tip of the S1 domain of the spike (S) protein, which is responsible for the high affinity binding of the virus to its receptor Angiotensin-Converting Enzyme-2 (ACE2), present on human epithelial and endothelial cells^8, 9^. For this reason, antibodies against the RBD that block its interaction with ACE2 are the most direct mechanism for neutralising the virus and indeed, one study reported that at least 90% of all SARS-CoV-2 neutralising antibodies in convalescent plasma were RBD reactive^8^. However, not all RBD-specific antibodies are capable of blocking the RBD-ACE2 interaction and neutralising virus infectivity. It is therefore important to determine not only whether an individual has antibodies against SARS-CoV-2 RBD, demonstrating a prior encounter with infectious virus or vaccination, but also whether they have NAb that are likely to protect from subsequent infection.

Plaque-reduction neutralization and SARS-CoV-2 microneutralisation assays are the gold standards for measuring virus neutralising antibodies^7^. These assays involve incubating SARS-CoV-2 with serial dilutions of serum and assessing residual infectivity in the sample. A major difficulty is that these assays require appropriate biocontainment laboratories, highly trained staff, and typically take several days for results^7^. Pseudovirus neutralisation assays are widely used alternatives that incorporate SARS-CoV-2 spike as the surface entry protein for a reporter virus into target that can be used in a BSL2/PC2 culture laboratory^10^. Another assay that has been developed for use in a PC2 laboratory is the plate-based ‘surrogate Virus Neutralisation Test’ (sVNT or C-PASS) that requires no cell culture^11^. This FDA-approved test relies on the ability of neutralising antibodies to bind to RBD and prevent its interaction with ACE2, thereby, inhibiting the signal which is measured by a colorimetric change in a cell-free ELISA-based assay. As the ability to block RBD-ACE2 binding is the major mechanism for neutralisation, the competitive ELISA is a surrogate for determining neutralising antibody activity^12^. This assay has been rapidly adopted around the world as a more reliable test for neutralisation than mere detection of anti-RBD antibodies, and several versions of this type of assay have now been published^13-15^. While the ability to run the assay in a PC2 laboratory in just a few hours is a major advantage compared to microneutralisation assays, nonetheless, it still has a number of drawbacks. In particular, it requires a venous blood sample and preparation of plasma, and transport to a dedicated laboratory with trained laboratory staff, which means long delays between testing and results. This delay may be particularly problematic in isolated areas or low-middle income countries where access to laboratories is often limited or impossible and reproducibility between labs may be an added challenge. A point-of-care test that uses the same principle of RBD-ACE2 inhibition to measure neutralising antibodies was recently published on a non-peer reviewed pre-print server^16^ but that test appears to be reliable only for relatively high levels of antibodies, and has not demonstrated the equivalence of plasma and whole blood that are essential for use at POC.

Our study describes the development of a prototype lateral flow POC assay, termed COVID-19 NAb-test™, that uses a similar principle to the sVNT/C-PASS assay, except signal is detected on a nitrocellulose test strip where ACE2 is immobilised and soluble RBD flows across the strip in the presence of patient sample. The RBD protein conjugated to colloidal gold leaves a visible red signal (test line) that directly reflects the amount of RBD bound to ACE2. Thus, in the presence of neutralising antibody, proportionately less (or no) RBD-gold will bind to ACE2, resulting in a fainter or absent test line. The assay is completed within 20-30 minutes and can be readily modified to include newly emerged variant RBDs (such as variants of concern [VOCs]) as required. The device also includes a second test line to detect the presence of total anti-RBD antibody (regardless of neutralisation activity), and a combined procedural control/reference line that allows visual estimation of neutralisation titre versus a pre-defined threshold. The test is also compatible with the commercially available Axxin AX-2XS reader, providing the potential for fully quantitative results and linkage to laboratory information systems. The COVID-19 NAb-test™ test provides a potential solution for POC testing of COVID-19 immunity that can be applied in many settings in both high- and low-middle income countries.

## Results

### Development of a lateral flow assay to measure RBD-ACE2 inhibition

The sVNT assay is a competition ELISA assay to measure the presence of neutralising antibodies in serum or plasma based on their ability to interfere with SARS-CoV-2 RBD binding to ACE2^11^. Using the same principle of ACE2-RBD binding interference, we developed a rapid, lateral flow sVNT assay to measure neutralising antibodies in biological samples at the point of care. We tested several different forms of RBD and ACE2 protein, with one or the other bound to the nitrocellulose strip that forms the stationary phase of the lateral flow assay. The approach ultimately adopted, because it provided the clearest signal and widest dynamic range, was to use a fusion protein of human ACE2 and human IgG1-Fc immobilised on the strip as the solid phase, and a complex of RBD-biotin bound to anti-biotin-gold (RBD-Au) as the mobile phase, passing over the immobilised ACE2-Fc through capillary action in the device flow (Figure 1A). After initial titration experiments (not shown), we settled on ACE2-Fc at 2mg/ml (0.5 µg of ACE2-Fc per 5 mm wide test strip) and detected a clear visible signal from binding of RBD-Au at 1 µg/ml RBD (Figure 1B). Titration of the RBD-biotin before mixing with this fixed amount of anti-biotin gold, yielded a clear visual signal over a range of concentrations from 0.06-2ug/ml (Figure 1C), with evidence of a “prozone” effect at the highest RBD-biotin concentrations and a linear titration at lower concentrations, allowing us to identify the appropriate level of RBD-biotin that would be readily titratable upon inhibition by neutralising antibodies. Importantly, the appropriate concentration of RBD-biotin must be determined for each batch of RBD-biotin to ensure assay performance. Using this format, we established an optimal concentration of 2μg/ml ACE2-Fc to coat the assay strip and a maximum of 1μg/ml RBD-biotin mixed with the anti-biotin gold. As a proof of principle, we tested the assay using four different anti-RBD mAb: two that were known to inhibit RBD-ACE2 binding (neutralising; clones 42 and 37) and two that were non-neutralising (clones CR3022^17^ and 4B). These results provided a clear dose response curve showing near complete inhibition of the visual ACE2-RBD-Au signal with the neutralising mAb while the non-neutralising mAb had no clear impact on the strength of the signal (Figure 1D). The intensities of the test lines were also quantitatively assessed on an Axxin AX-2X strip reader and a clear dose-dependent decline in signal strength was observed with increasing amounts of the neutralising mAb (Figure 1E).

**Figure 1.**
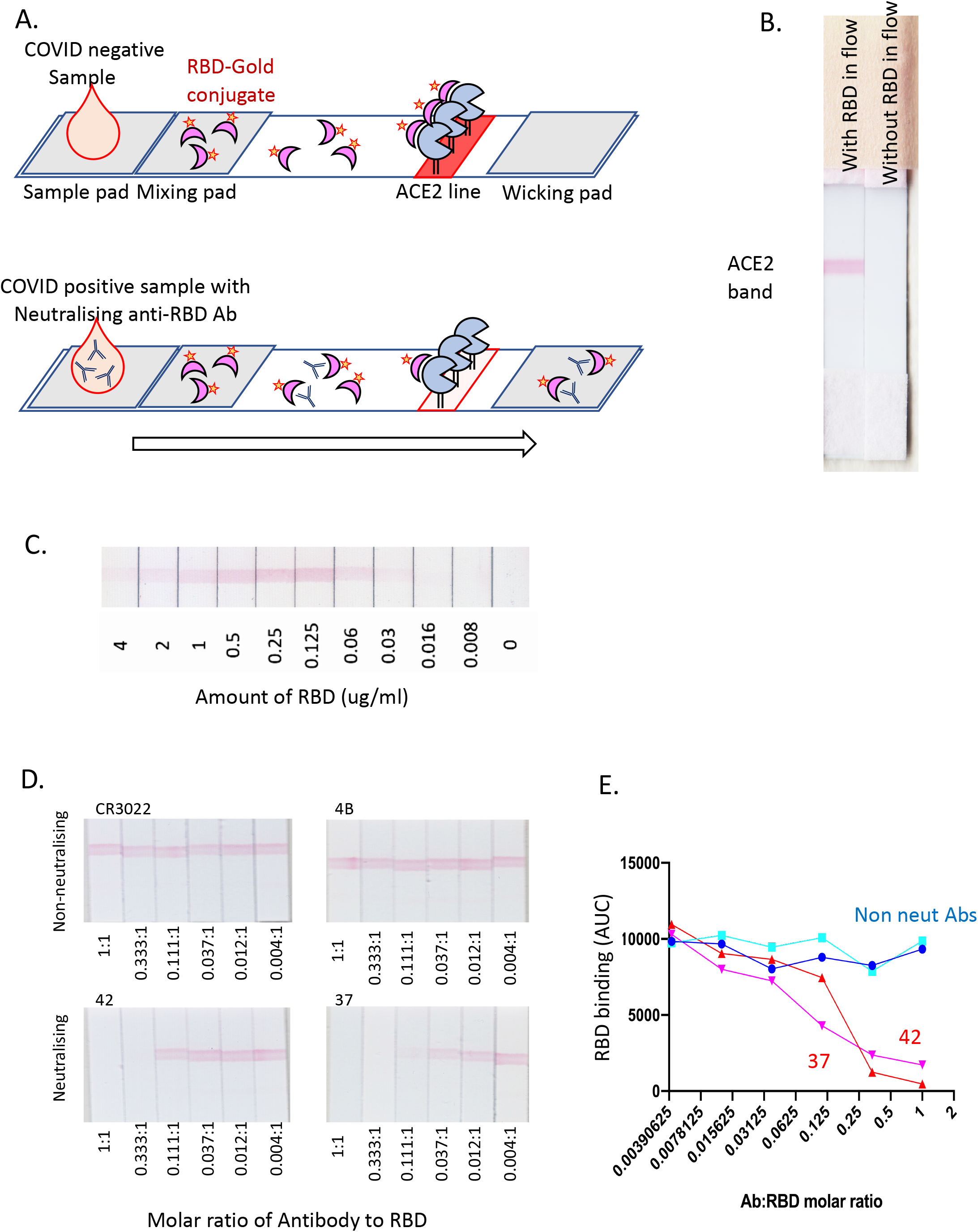
Development of a lateral flow assay to detect NAb. **A**. A schematic depicting our lateral flow COVID-19 NAb assay design. **B**. A photograph of two lateral flow strips side by side, with the left one showing a red band when RBD-Au has been included in the flow. **C**. Titration of RBD-Au run at a range of concentrations across the strips. **D**. pre-incubation of non-neutralising (CR3022, 4B) and neutralising (42, 37) anti-RBD-specific monoclonal antibodies at molar ratios as depicted. **E**. Graph represents intensity of red lines from Axxin strip reader.

### Testing COVID-19 patient samples

Having established that the assay can detect the presence of neutralising antibodies, we tested 79 COVID-19 patient plasma samples and 47 COVID-19 negative control samples, as well as samples from patients with other virus infections (Figure 2)^18, 19^. As expected, the pre-COVID-19 healthy control samples showed minimal fluctuation in the intensity of the RBD-ACE2 signal, within a range of ±20% around the mean (Figure 2A). In contrast, COVID-19 patient samples showed a broad range of inhibition in the intensity of the RBD-ACE2 test line, with some samples almost completely inhibiting the signal (Figure 2A). These samples were measured using the Axxin strip reader and the % inhibition within the COVID-19+ samples relative to the COVID-19-samples was calculated as: % inhibition = (1 - sample intensity/median intensity of COVID-19-ve samples) x100. Samples taken from patients with other virus infections including RSV, influenza and picornaviruses all showed no inhibition greater than 20% compared to control COVID-19 negative samples (Figure 2B). We plotted these inhibition values against the neutralising titres from the same samples determined using a SARS-CoV-2 microneutralisation assay, revealing a strong correlation in neutralisation (R^2^=0.72, p<0.0001) (Figure 2C). Furthermore, our assay has the sensitivity and dynamic range to distinguish between strong, weak and absent neutralising antibody levels.

**Figure 2.**
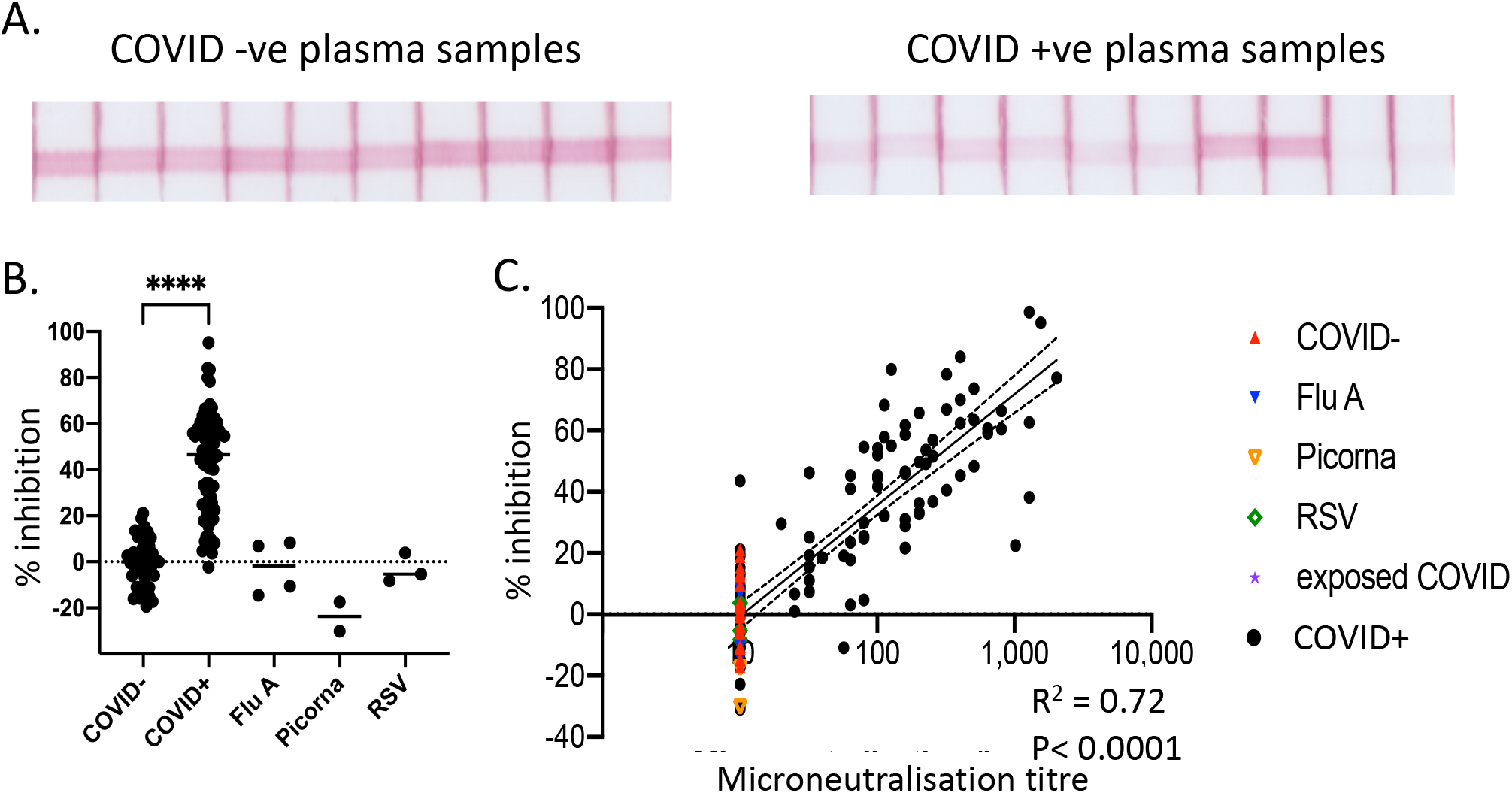
Testing the lateral flow assay on COVID- and COVID+ plasma samples. **A**. Ten representative COVID-19 -ve and COVID-19 +ve plasma samples. **B**. Tests of 47 COVID-19 -ve, 79 COVID-19 +ve, and 9 non-COVID virally infected individuals were converted from stripe intensity from each sample to a % inhibition based on the formula % inhibition = (1 - sample intensity/ median intensity of COVID-19-ve samples) x100. Plotted as % inhibition on y axis. **C**. Readings from lateral flow strips were plotted against readings for same samples derived from virus microneutralisation assay.

### Development and testing of a prototype assay for POC use

To facilitate intended use of the test in a POC setting, the assay strip configuration was modified to allow the use of a self-contained cartridge (Figure 3). In this configuration, a small volume of plasma is added to well A, where the sample rehydrates the RBD-Au complex and begins to migrate into the mixing pad. Three drops of running buffer (buffered saline) are then added to well B, and the buffer chases the plasma and RBD-Au complex mixture along the length of the test strip within the closed cassette (Figure 3A and B). The sample pad also contains a defined amount of gold-conjugated chicken IgY, which serves both as an assay running control and internal reference material. The sample then reaches the immobilised ACE2-Fc line, where the amount of RBD-gold complex binding will be inversely related to the amount of neutralising antibody in the sample. Two additional lines have been incorporated on these strips. The reference line is a stripe of anti-chicken IgY that binds the chicken IgY-gold, serving as a control to show that the test has run successfully and also providing a reference line for comparison to the ACE2-RBD-Fc test line for both semi-quantitative and quantitative measurement of neutralising antibody. For visual (semi-quantitative) estimation, the reference line represents the intensity associated with a predetermined amount of inhibition (eg. 50% as used in these examples). The intensity of the line can be adjusted at manufacture by varying the amount of anti-chicken IgY and/or chicken IgY-gold that are added, allowing a direct visual comparison between the test and reference lines to assess whether a sample has sufficient levels of neutralising antibody. This approach is similar to the visual reference lines incorporated in the POC Visitect® CD4 T-cell tests that have cutoffs of either 350 or 200 CD4 T-cells/µl (Omega Diagnostics, UK)^20^. For quantitative estimation (using the Axxin strip reader), the % inhibition for samples is calculated as: % inhibition = (1 – (test line intensity/[2x reference line intensity])) x100. The third line is striped with RBD protein, which forms a double-antigen sandwich with RBD-gold and anti-RBD antibody derived from the sample being tested. This line therefore indicates the presence of total anti-RBD antibody, regardless of whether or not it is neutralising and capable of blocking RBD-ACE2 interaction, and is analogous to many total antibody tests currently available for SARS-CoV-2. Our prototype assay was used to examine 72 samples from healthy pre-COVID controls and 200 samples from COVID-19 patients, examples of which are shown in (Figure 3C). The data showed a high degree of consistency across the control samples and a wide range of inhibition from 0-95% in COVID-19 patient samples (Figure 3D) calculated using the formula above. When the data were plotted against neutralising titre data from the same samples determined by microneutralisation assay, a clear correlation was observed (R^2^ = 0.69, P<0.0001) (Figure 3E).

**Figure 3.**
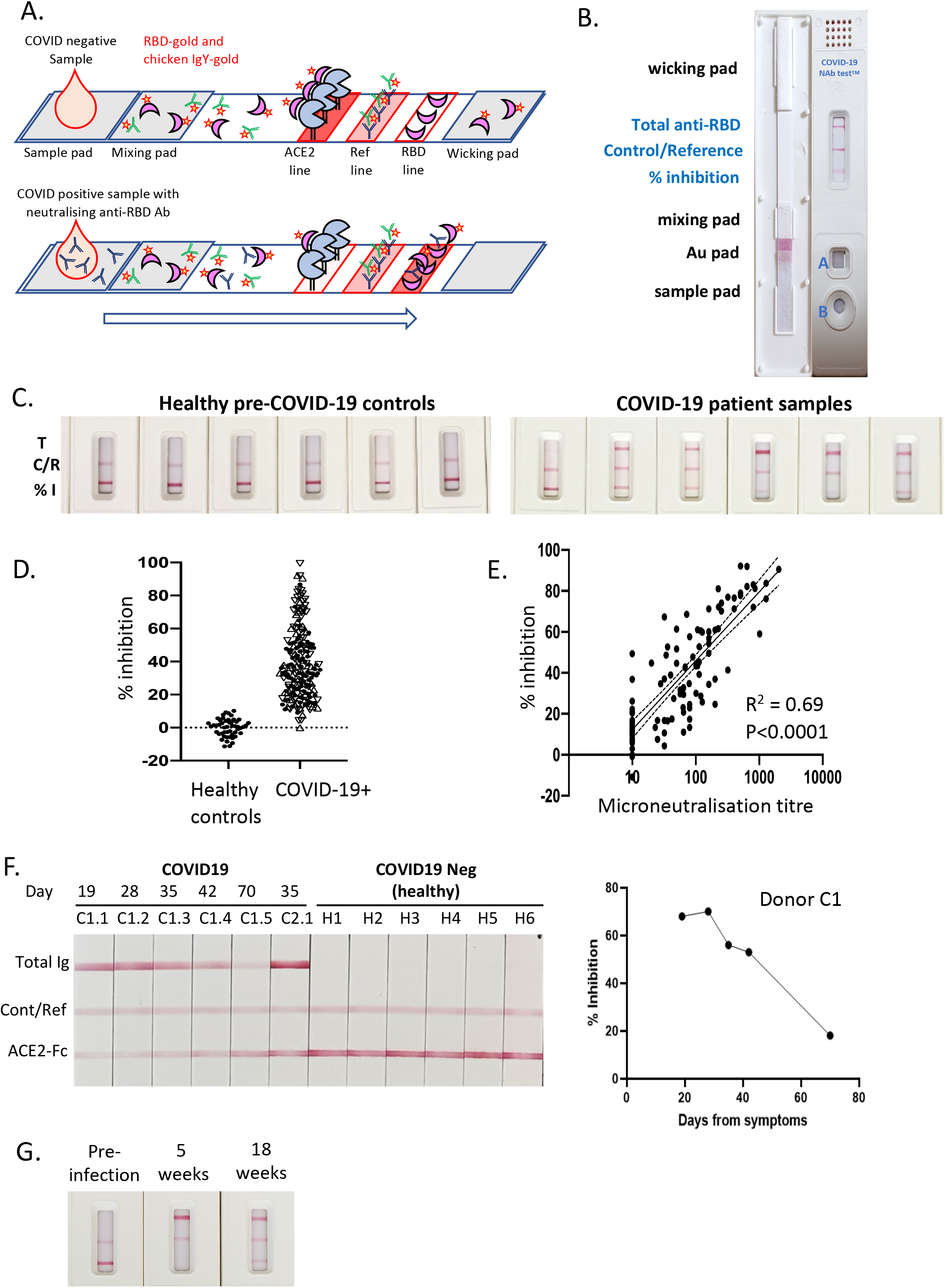
Development of a prototype test cassette for measuring Nab against SARS-CoV-2. **A. and B**. A schematic and a photograph depicting our prototype lateral flow assay cartridge design. **C**. 6 representative healthy pre-COVID-19 samples and 6 representative COVID-19 patient samples as run on the prototype lateral flow cartridge. **D**. Conversion of stripe intensity from each sample to a % inhibition based on the formula % inhibition = (1 - sample intensity/ median intensity of COVID-19-ve samples) x100. **E**. Readings from lateral flow strips were plotted against readings for same samples derived from virus microneutralisation assay. **F**. Samples from an individual that were collected from 3 to 10 weeks. Six COVID-19 -ve samples were also run for comparison. Results from Donor C1 are plotted as %inhibition vs days from symptom onset. **G**. Pre- and post-infection samples from a symptomatic patient at indicated timepoints.

Previous reports have suggested that neutralising antibody levels may decrease quite rapidly, within weeks to months of infection^21-23^. Accordingly, tests that can measure neutralising antibody may be useful to confirm adequate levels of neutralising antibody at regular intervals after infection or vaccination. We examined a series of longitudinal samples from a single subject (C1) beginning 3 weeks after PCR-confirmed diagnosis of infection with SARS-CoV-2 (Figure 3F). Over the period of 3-10 weeks, the degree of ACE2-RBD inhibition shown in the test line can be visually observed to decline in the images of the strips, and using the Axxin reader it is shown to decline from ∼70% to 18%, in parallel with declining levels of total anti-RBD Ig. In contrast, a single sample from subject C2.1 shows high levels of total anti-RBD Ig but negligible (3%) ACE2-RBD inhibition (Figure 3F). This suggests that this subject had limited neutralising antibody titres despite the presence of measurable total anti-RBD antibodies, although it is possible that this subject had other neutralising antibody specificities that do not target the RBD-ACE-2 interaction. The six healthy controls used in this experiment (H1-H6) showed no anti-RBD antibody and no detectable inhibition. Next, we examined serum samples from an individual with symptomatic, PCR-confirmed SARS-CoV-2 infection where a pre-infection sample was also available, which formally shows that a response was not detectable pre-infection (Figure 3G). The sample taken 5 weeks after symptom onset showed seroconversion for anti-RBD and very strong RBD-ACE2 inhibition, with some decline in anti-RBD and RBD-ACE2 inhibition observed at the 18-week timepoint.

### Serum samples from SARS-CoV-2 immunised macaques

Next we carried out longitudinal tests samples from macaques that had been immunised with experimental SARS-CoV-2 spike vaccines that induced high titre NAb responses^24^. Results in Figure 4A are shown from 3 macaques primed with spike protein vaccine with Addavax adjuvant, and boosted with soluble RBD protein on day 21, and from 8 additional macaques (Figure 4B) primed with whole spike protein (day 0) and boosted with spike protein (day 21), as described^24^. Blood samples were taken prior to the prime (day 0), post-prime (day 14; five animals only), prior to the boost (day 21), and post-boost (day 42). Graphs depict from left to right: raw Axxin readings; % inhibition (relative to pre-bleed baseline for each animal); and total anti-RBD results (Figures 4A and B). While some evidence of RBD-ACE2 inhibition was observed with the post-prime/pre-boost samples (Figures 4A and B), the post-boost (day 42) samples all showed clear RBD-ACE2 inhibition ranging from 50% to 95%. The total anti-RBD line also became more prominent in all animals following the boost (Figure 4A and B). Given that NAb responses are induced by most current SARS-CoV-2 vaccines, we anticipate that similar results will be derived from humans that are immunised with SARS-CoV-2 vaccines, and these studies demonstrate that this assay can provide clear evidence of total anti-RBD antibodies and neutralising antibodies from vaccine recipients. It is also noteworthy that these data demonstrate that our assay works with non-human samples and may also be valuable for assessing SARS-CoV-2 immunity in animal models and SARS-CoV-2 exposure in other species.

**Figure 4.**
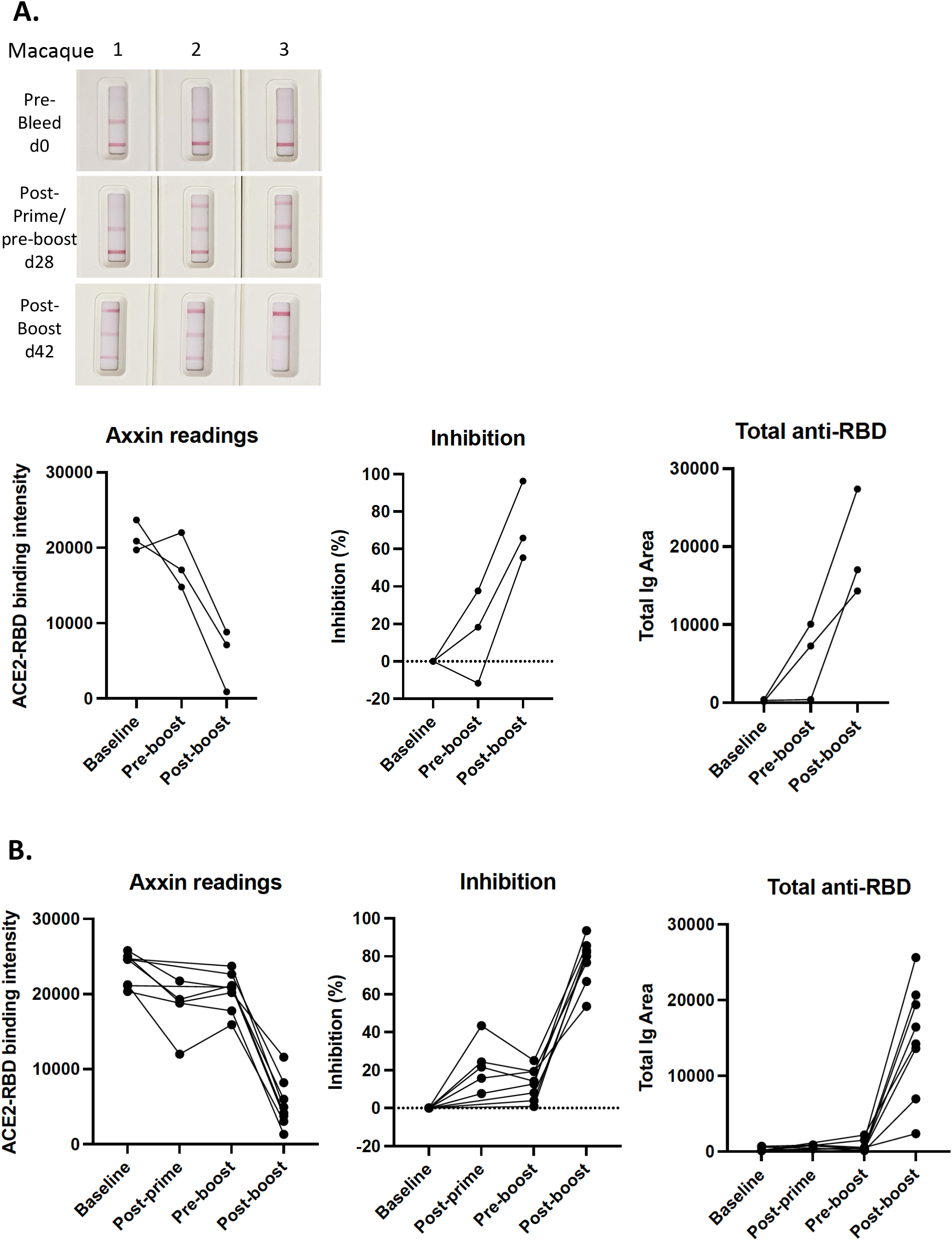
Analysis of plasma samples from immunised macaques. **A**. Representative images of test results from longitudinal samples from macaques immunised with SARS-CoV-2 spike protein (prime), RBD protein (boost) (left) and data plotted to compare % inhibition or anti-RBD antibody shown at baseline (day 0), pre-boost (day 28) and post-boost (day 42). Graphs depict raw Axxin data readings linked for each animal (left); % inhibition relative to baseline reading linked for each animal (middle); and total anti-RBD reading linked for each animal (right). **B**. Samples from a different cohort of macaques immunised with SARS-CoV-2 spike protein (prime), and spike protein (boost), shown at baseline (day 0), post-prime (day 14), pre-boost (day 28) and post-boost (day 42). Graphs depict data as described in A. Samples from macaques were derived as previously described^24^.

### Neutralisation testing for viral variants

In addition to waning levels of antibody over time, the long-term efficacy of antibody-based immunity to COVID-19 from vaccination or prior infection is being challenged by the emergence of variants of concern (VOC), with single and multiple mutations in the spike protein and especially in RBD that may affect binding affinity to ACE2, transmissibility (R0) and/or susceptibility to neutralising antibodies. Testing of immunity may therefore require consideration of circulating VOCs. Our test was designed to facilitate the substitution of variant RBD proteins by the use of RBD-biotin and anti-biotin gold nanoparticles, and we evaluated four different versions of RBD-biotin in our assay (Figure 5), choosing variants observed during second wave infections in June 2020 that have been reported to have varying sensitivity to some mAbs^25, 26^. The original (Wuhan) RBD sequence and variants S477N, S477I and N439K were expressed with biotin tags, purified and titrated before mixing with a fixed amount of anti-biotin gold (Figure 5). Each of the four RBDs showed essentially identical titration curves for binding of RBD-biotin-Au complexes to ACE2 (Figure 5A). We next assessed the susceptibility of each variant to inhibition by patient antibodies, again using longitudinal plasma samples from patient C1 (infected in February 2020). As shown in Figure 5B, the level of inhibition for all variants was around 90% at week 3 after symptom onset, reducing to around 80% at 5 and 50% at 10 weeks for each variant, with only some subtle differences. The level of total antibody measured with each variant RBD-biotin-Au was also similar at each time. While these variants did not reveal any striking differences, these experiments demonstrate the utility of our assay for the detection and assessment of neutralising antibody responses to RBDs from variants of SARS-CoV-2 through substitutions of the corresponding recombinant RBD-biotin forms.

**Figure 5.**
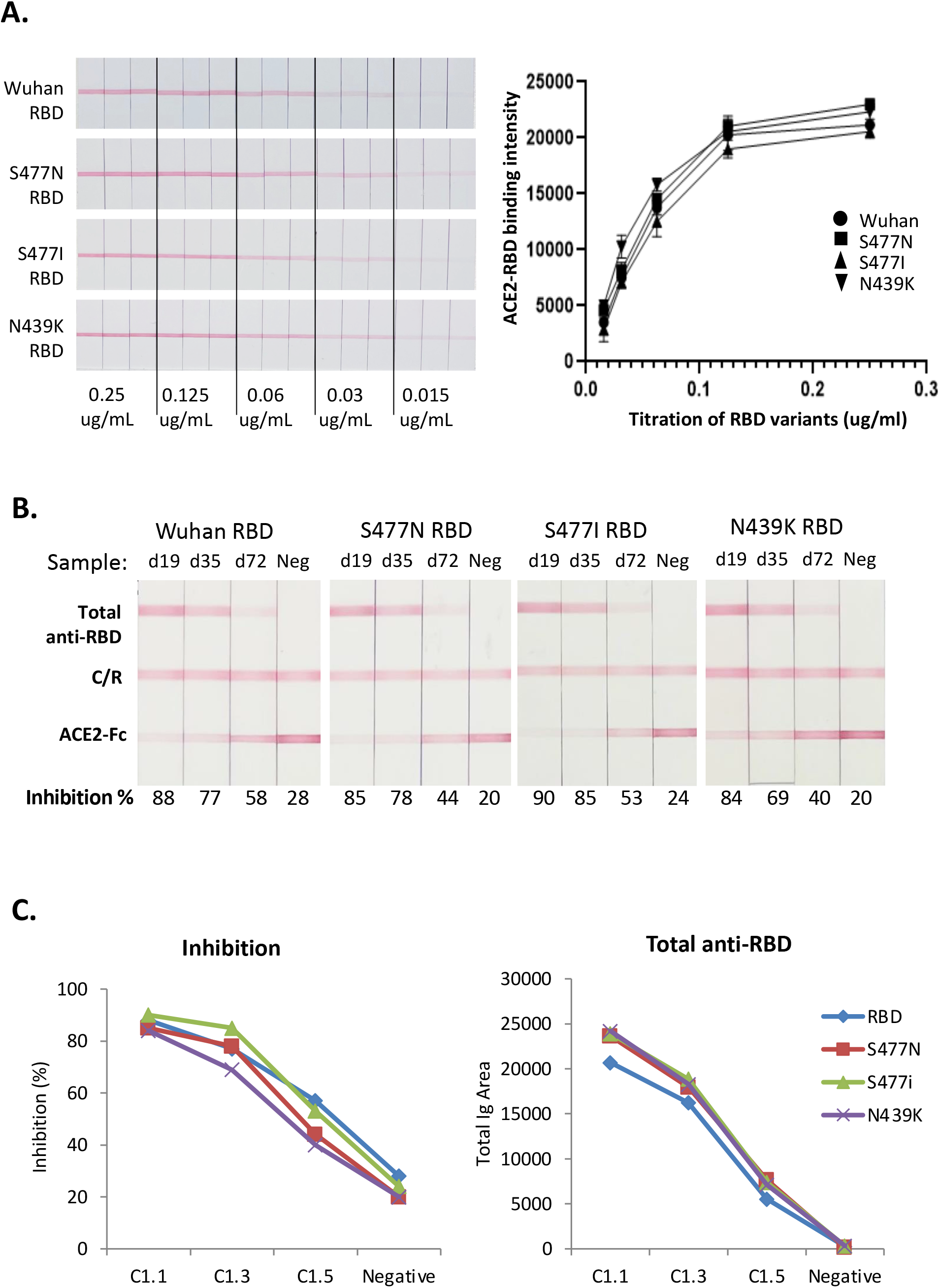
The test cassette can be adapted to work with variant RBDs. **A**. Original Wuhan RBD and 3 variant RBD (S477N; S477I; N439K) gold conjugates were titrated on ACE2 strips, in triplicate. Red line depicts direct binding between RBD and ACE2. Each variant showed a similar titration curve. **B**. Longitudinal samples from donor C1 were titrated against strips in the presence of Wuhan RBD or the 3 variants. **C**. Data from B plotted to compare % inhibition, relative to the reference line, against each different RBD or anti-RBD antibody.

### Compatibility with whole blood for use at point of care

Lateral flow tests are suitable for use at POC, but for blood-based tests they must function with the use of whole blood rather than plasma or serum. To achieve this, we further modified the test by addition of anti-glycophorin A antibodies to the sample pad along with the RBD-Au complexes. Anti-glycophorin A agglutinates red blood cells, while white blood cells, plasma and the RBD-Au complexes are able to flow as normal; the white blood cells are subsequently retarded at the interface of the nitrocellulose membrane^27^. We also used a single well cartridge as we found this gave better separation from the trapped red blood cells to the test area of the strip (Figure 6A). Figure 6B shows the results of test strips using whole blood from a COVID-negative donor, spiked with varying levels of neutralising mAb#42. This showed good correlation of ACE2-RBD inhibition versus mAb concentration, providing proof of concept for the compatibility of the test format with whole blood. We noted that the total anti-RBD antibody line was fainter than expected with these samples, probably a consequence of a single specificity mAb compared to polyclonal antibodies, including various isotypes, present in plasma samples. To provide further proof of concept for using whole blood versus plasma, longitudinal samples from subject C1 were tested on the device strips shown in Figure 6A using either 15 µl of plasma, or 15 µl of plasma mixed with 15 µl of packed red blood cells from a COVID-19 negative donor, emulating 30 µl of a whole blood sample with 50% hematocrit (Figure 6C). The visual appearance and quantitative readout of the plasma and reconstituted whole blood samples were very similar for both the level of inhibition and the level of total anti-RBD (Figure 6C), which suggests that the COVID-19 NAb-test™ test should have utility for true POC use.

**Figure 6.**
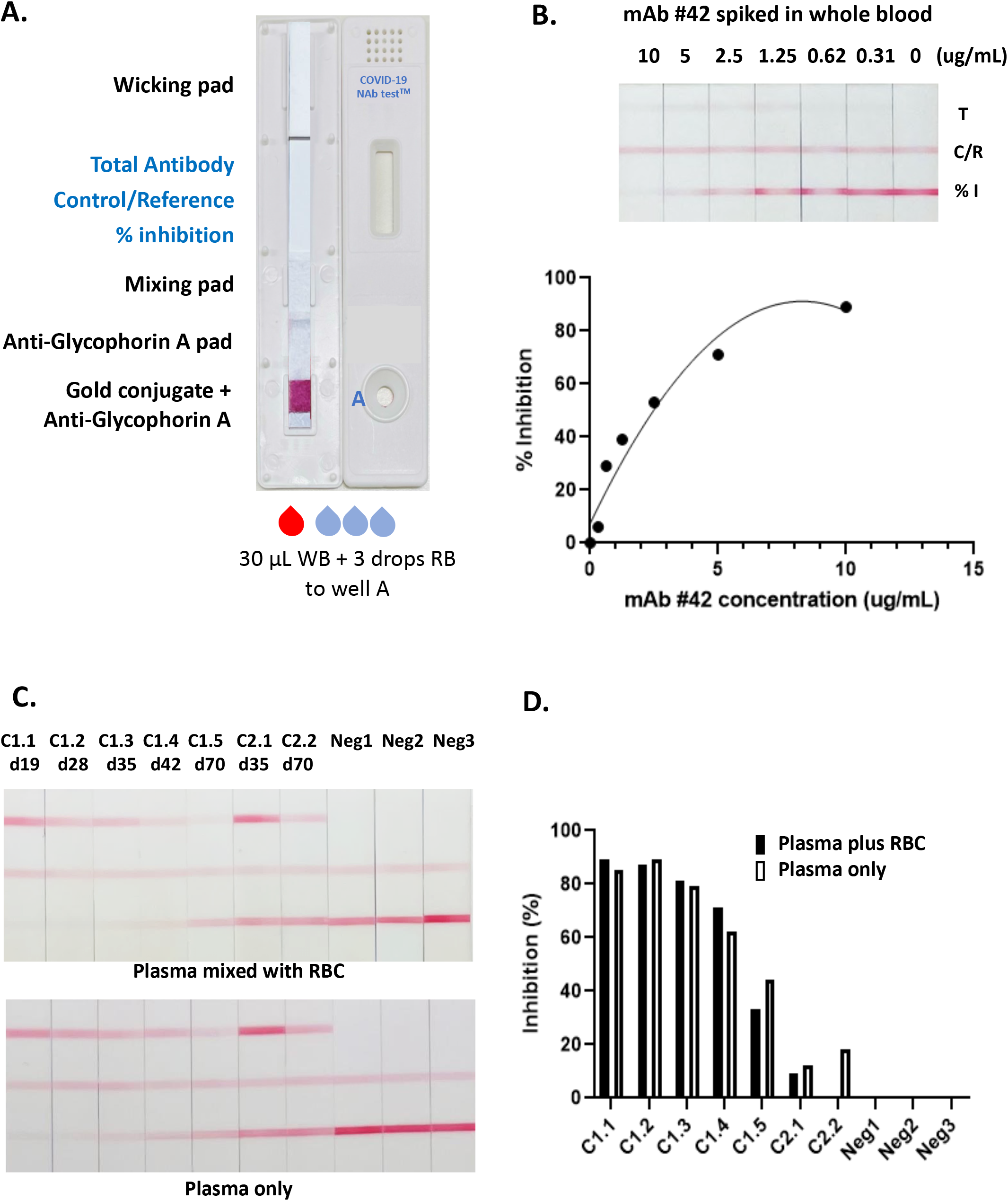
Development of a test cassette that measures NAb from whole blood. **A**. An image of the cartridge used for testing whole blood, incorporating anti-glycophorin-A pad, and a single well for sample loading. **B**. Whole blood was spiked with varying concentrations of neutralising mAb 42 and tested on strips incorporating anti-glycophorin A which captures red blood cells. Graph depicting %inhibition (relative to the reference line, from Axxin strip reader) versus amount of mAb 42 spiked into sample. **C**. Longitudinal plasma samples from donor C1 were either reconstituted with an equal volume of packed red blood cells and run as 30 µl whole blood, or run as 15 µl plasma only. Two longitudinal samples from donor C2 and three COVID-19 negative samples also shown. **D**. Graph depicting % inhibition (relative to the reference line, from Axxin strip reader) from data shown in C, comparing plasma added to red blood cells, or plasma only.

### Confirmation of immunity in immunised humans

To validate our test with a sample of post-(Pfizer) vaccination humans, we tested a cohort of vaccinees at least 2 weeks following the second (boost) dose, compared to a group of unvaccinated individuals (red), using the whole blood test (Figure 7A and B). There was a clear correlation (R2 = 0.75, p<0.0001) between the results from our assay and microneutralisation, where >50% inhibition at the RBD-ACE2 test line was exhibited by 26 out of 27 samples with a microneutralisation titre of greater than 40, which we consider represents a reasonable lower limit for immunity against SARS-CoV-2^6^. There was one outlier in the cohort of post-boost individuals with microneutralisation titre of 157 that failed to show inhibition in our RBD-ACE2 test line, despite showing a clear reading for total anti-RBD antibody (Figure 7B). This sample may reflect an individual who generated neutralising antibodies that target parts of the spike other than the RBD.

**Figure 7.**
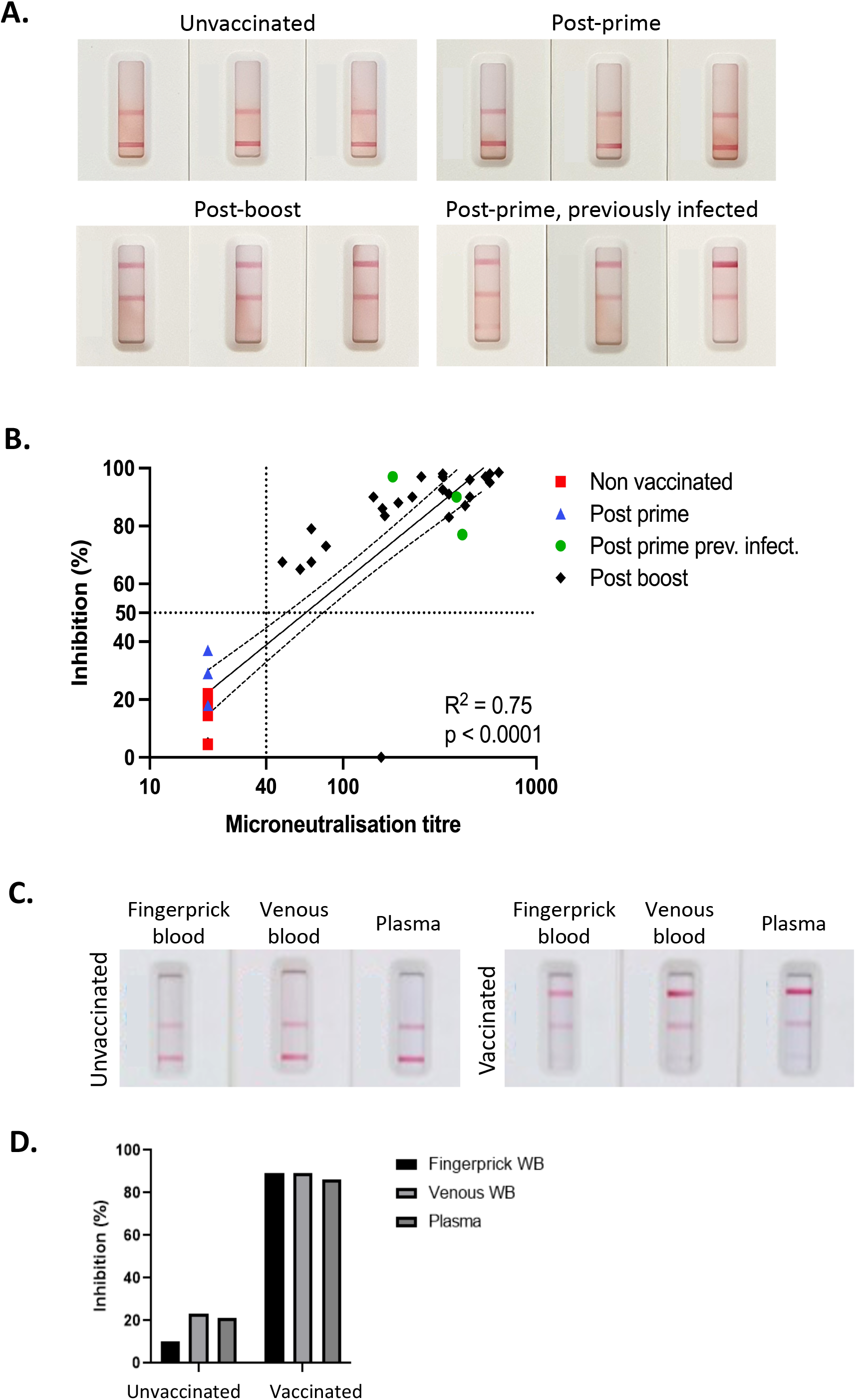
Assessment of NAb in post-vaccinee samples using whole blood. **A**. Representative images of test results from human venous blood samples from (top left) unvaccinated, (top right) post-prime vaccinated (Pfizer, mRNA spike vaccine), (lower left) post-boost vaccinated and (lower right) post-prime vaccinated following previous infection with SARS-CoV-2. B. Inhibition readings from the Axxin strip reader from samples from groups described in A. were plotted against readings for the same samples derived from virus microneutralisation assays. C. Samples from fingerprick whole blood were compared to venous whole blood and plasma from two individuals, one unimmunised and uninfected and one previously primed and boosted with the Pfizer vaccine (22 days post-boost). D. Inhibition readings from the Axxin strip reader are depicted in the graph.

Six samples from vaccinees after a single dose were also included in this study (Figure 7A and B). Three of these samples (blue) showed little or no inhibition of RBD-ACE2 binding in our test, and no microneutralisation activity above the background titre of 20 and were thus very similar to unvaccinated controls. This reflects the fact that naïve individuals require a prime and boost to develop a strong neutralising antibody response. Interestingly, however, the other three post-prime samples (green) showed strong inhibition of RBD-ACE2 binding in our test, which correlated with strong microneutralisation titres. This group were from a study of people who had previously been infected with SARS-CoV-2 and have subsequently been immunised. This highlights the ability of our test to detect NAb regardless of whether they are induced by infection, or vaccination, or a combination thereof.

Lastly, to ensure that fingerprick-derived whole blood works as effectively as venous whole blood, we compared fingerprick-derived whole blood versus venous derived whole blood versus plasma from samples derived from an unvaccinated individual and a vaccinated individual (Figure 7C). These data demonstrate that fingerprick derived whole blood provides similar results to venous whole blood and plasma, in our assay, confirming that this test can be easily used at point of care with a simple finger prick blood sample.

## Discussion

As an increasing number of approved COVID-19 vaccines are now being rolled out around the globe, the hope is that individuals who have developed immunity will contribute to population-level herd immunity, and facilitate a return to a pre-COVID-19-like existence. However, we currently lack widely-deployable tools to answer the critical questions of who is actually immune, and for how long immunity lasts, and whether their immunity will protect them from new variants of SARS-CoV-2. Tests that detect viral RNA do not provide this information, nor do tests that measure total anti-virus antibody, whether they are ELISA or lateral flow-based. As we and others have shown, the existence of antibodies that can bind to viral proteins, including RBD-specific antibodies, does not necessarily demonstrate the existence of neutralising antibodies. It is generally accepted that to be protective, neutralising antibodies need to target the spike protein, and in SARS-CoV-1, antibodies against other structural proteins (E, M and N) were unable to mediate protective immunity^28^. Furthermore, at least 90% of neutralising activity is focussed on the RBD of the spike protein, and functions predominantly by preventing interaction with cellular receptor ACE2^8^.

It is not yet clear what titre of neutralising antibody is adequate for protection against SARS-CoV-2. Studies in macaques involving transfer of SARS-CoV-2 neutralising antibody from immune to naïve animals suggest that NAb titres of approximately 50 (based on pseudovirus assay) or total anti-RBD ELISA titres of approximately 100 are protective against infection, although the assessment was based on a limited number of animals^29^. Furthermore, several studies have shown that neutralising antibody titres following infection wane with time, which means regular testing for immunity will likely be required. Published estimates vary, but most studies agree that neutralising antibody levels begin to decline within a few weeks of infection. One study of 149 individuals following COVID-19 disease showed that one third of these individuals had neutralising titres below 1:50 by roughly 39 days after symptom onset^30^. Depending on the maximum antibody titre, which is typically linked to the severity of infection, it may take between 4 months and 1 year for the level of neutralising antibody to drop below detection limits^31^.

Scenarios such as national or international travel and border control, verification of vaccination status, risk stratification of quarantine requirements, screening of donors for convalescent plasma or antibody therapy, or returning to work or school, may require regular assessment of immune status. A rapid (20 minute) test using fingerprick whole blood overcomes limitations associated with current tests, requiring venous blood draw and laboratory equipment with results taking a minimum of three hours. Portable strip-reading instruments such as the Axxin AX-2XS used here are commercially available and can be deployed in non-laboratory locations such as airports and aged care or educational facilities, and the use of dedicated readers (and potentially even smartphone Apps) can provide a quantitative assessment of neutralising antibody levels. Even in the absence of a dedicated instrument, a semi-quantitative assessment by eye, visually comparing the intensity of the test line to the reference line, should provide sufficient information on whether an individual has an acceptable level of immunity, using the same principle used to determine CD4 T-cell levels in the semi-quantitative point of care VISITECT® CD4 tests^20^.

An additional risk that threatens to reduce the durability of immune protection is posed by the emergence of mutant strains of SARS-CoV-2, including the B.1.1.7 lineage (also known as the UK variant), the B.1.351 lineage (South Africa variant) and B.1.1.28 lineage (Brazil) that are considered to be variants of concern (VOCs). Each of these VOCs are more infectious, and potentially induce more severe disease. While they carry mutations throughout various viral proteins, the key mutations appear to be those in the RBD region (N501Y in B.1.1.7; N501Y, E484K and K417N in B.1.351; and N501Y, E484K and K417T in B.1.1.28) that markedly increase ACE2 binding affinity of the virus (N501Y) or impact recognition by antibody (E484K;K417N). Several studies have shown that these variants, particularly B.1.351 and B1.1.28, are far more difficult to neutralise, effectively lowering neutralising antibody titres by up to 33-fold in convalescent plasma and 8.3 fold in individuals vaccinated with the Pfizer and Moderna vaccines^5^. Neutralising antibody assays will need to incorporate these variants to accurately determine immunity based on the currently and locally circulating strains, helping to guide strategies for boosters to address waning immunity and/or VOCs. The ability to interchange RBDs carrying relevant mutations as required (Figure 5), further highlights an important advantage of our assay, especially compared to the measurement of total S- or RBD-specific antibody or IgG which is unlikely to reflect neutralisation escape. Moreover, the technology could theoretically be adapted in future iterations to simultaneously measure against multiple mutants by using multiple parallel strips. Our longitudinal analysis of serum samples from macaques that had been immunised with experimental spike protein vaccines, as well as whole blood samples from humans immunised with spike mRNA vaccines, demonstrated that our assay provides a clear readout of anti-RBD antibody as well as neutralising antibody following immunisation (Figures 4 and 7).

Limitations of this study include the underlying premise that we will need to test people from time to time to assess their antibody status and risk of infection. While this is likely to be valuable, it should be acknowledged that even if/when antibody titres fall, it is possible that memory B and T cells will provide rapid and effective protection upon re-exposure. Therefore, while antibody titres may decline, that does not necessarily mean that the individual is fully susceptible to disease. Indeed, repeat testing with an easy and rapid POC test has the potential to exaggerate fear of re-infection risk. Furthermore, it remains to be determined what the policy response will be for individuals showing low to undetectable Ab, although in the case of hepatitis B virus immunisation for high-risk workers, repeated booster immunisations are sometimes recommended for those who do not show clear seroconversion. Lastly, while 90% of neutralising antibodies are directed toward the RBD, inhibiting its binding to ACE2, it is possible that neutralising antibodies against other parts of the spike protein may be prominent in some individuals, yielding a false negative or under-estimate of immunity using our assay, and indeed we observed one such sample in our study of vaccinee samples. Conversely, it may be reasonable to assume that RBD-specific immunity measured with the COVID-19 NAb-test™ will represent a minimum estimate of immunity to the variant included in a particular version of the test.

SARS-CoV-2 continues to spread and threatens to disrupt human activity for years to come. Fortunately, we already have vaccines that are proving to be effective at reducing disease that are now being rolled out in many countries, but countering that, many countries have low vaccine coverage and ongoing high rates of infection, raising the potential for immune-mediated selection of escape variants. It is possible that immunity tests may become a normal part of life, at least in some circumstances such as international travel, as we adjust to life with SARS-CoV-2 as an endemic virus. Our test should contribute to more widespread access to immunity testing for many populations, including those in resource-poor areas with limited access to laboratories and equipment for more conventional virus neutralisation tests or ELISA-based inhibition assays.

## Materials and Methods

### Human Samples

All human specimen materials were considered infectious and hazardous and handled using standard biosafety procedures. Plasma: Blood specimen was collected in a lavender, blue or green top Vacutainer® collection tube containing EDTA, citrate or sodium heparin (respectively), using venepuncture. Plasma was separated by centrifugation. Plasma was then carefully withdrawn and decanted into a new pre-labelled tube. Serum: Blood specimen was collected in a red top Vacutainer® collection tube without the presence of coagulants. Blood was allowed to clot, and the serum separated using centrifugation. The serum was then carefully withdrawn and decanted into a new pre-labelled tube. Specimens were frozen at −20°C for longer term storage and tested as soon as possible after thawing (up to 3 days). For frozen samples, more than 4 freeze-thaw cycles were avoided. Prior to testing, frozen specimens were brought to room temperature slowly and gently mixed. Samples containing visible particulate matter was clarified by centrifugation before testing. Samples demonstrating gross lipaemia, gross haemolysis or turbidity were discarded to avoid interference on result interpretation. Whole blood: Samples of blood were obtained by venepuncture (stored for up to 24 hours at 2-8°C as required), or fingerprick (used immediately).

Human experimental work was conducted according to the Declaration of Helsinki Principles and the Australian National Health and Medical Research Council Code of Practice. All participants provided written informed consent prior to the study. The study was approved by the Alfred Hospital (#280/14), The University of Melbourne (#2056761, #2056689, #1442952, #1955465, #202121198153983 and #2020-20782-12450-1), Melbourne Health (HREC/68355/MH-2020, HREC/66341/MH-2020) and James Cook University (H7886) Human Research Ethics Committees. Plasma samples including COVID-19 acute/convalescent and vaccinated samples, were obtained from: The Alfred Hospital, University of Melbourne, James Cook University, The Royal Melbourne Hospital. Some samples obtained from The Royal Melbourne Hospital Department of Microbiology were part of a previously published patient cohort^4^ and patients C1, C2 and COVID-negative blood and plasma from the Burnet Institute. The vaccinee samples are collected under ethics number 2021-21198-15398-3 through the University of Melbourne. Samples were collected by venepuncture into heparin tubes. Whole blood was provided from those tubes, and plasma was collected after centrifugation. All vaccinees reported no prior positive tests for SARS-CoV-2 infection. Unless otherwise stated, samples were collected at day 14 (plus or minus 1 day) after administration of the 2^nd^ vaccine dose.

### Macaque samples

Macaque serum samples were derived from stored samples from macaques that had been immunised with SARS-CoV-2 spike or RBD proteins, as described in detail in a recent study^24^. Animal studies and related experimental procedures were approved by the University of Melbourne Animal Ethics Committee (no. 1714193, no. 1914874). Macaque studies and related experimental procedures were approved by the Monash University Animal Ethics Committee (no. 23997).

### Reagents

Double stranded DNA encoding the truncated SARS-CoV-2 RBD (N334-P527) with C-terminal AVI-tag was synthesised (IDT) and cloned into the pHLSec expression vector. The protein was expression by transient transfection of Expi293F cells using Expifectamine transfection kits (ThermoFisher Scientific)^32^. Expressions were harvested on day 6, and the protein purified by Ni-NTA. SARS-CoV-2 RBD-avi protein was then enzymatically biotinylated using recombinant BirA protein. Biotinylated SARS-CoV-2 RBD-biotin was further purified by size-exclusion chromatography using a HiLoad Superdex 75 16/600 (Cytiva). Biotinylation was found to be at least 95% efficient by streptavidin gel-shift.

Variants (N439K, S477N/I, E484K, N501Y, K417T) of SARS-CoV-2 RBD were expressed in Expi-293F cells stably transfected with BirA ligase at 34C and supplemented with L-Biotin. Four days after transfection tissue culture supernatants were clarified and target proteins were purified by IMAC using Talon metal affinity resin (Clontech Laboratories) following the manufacturer’s recommendations. The eluted proteins were subject to gel filtration using a Superdex 200 16/600 column (GE Healthcare) with PBS as the liquid phase. Fractions corresponding to monomeric RBD were pooled and concentrated in Amicon Ultra 30kDa devices (Merck) prior to use.

Double stranded DNA encoding the truncated extracellular domain of human ACE2 (aa Ser19-Ser740, accession BAB40370) was fused by a Gly-Gly-Gly-Gly-Ser linking sequence to human IgG1-Fc region (Thr223-Lys447; Eu numbering, Accession no. AXN93652.1), was expressed in Expi293F cells as above, and purified using Protein A sepharose (Cytiva) followed by size-exclusion chromatography using a HiLoad Superdex 200 16/600 (Cytiva).

Anti-RBD mAb were kindly provided by Dr Adam Wheatley; anti-biotin-gold (40 nm, BBISolutions, UK); anti-glycophorin A (Epiclone, Seqiris, Australia).

### Equipment

The Axxin AX-2XS instrument reader was obtained from Axxin, Melbourne, Australia.

### Production of Lateral flow device

The device cassette consists of a plastic housing (Nanjing BioPoint Diagnsostics, PR China) with loading wells and a window to read results. Within the cassette is a nitrocellulose membrane strip (Sartorius, Germany), with, at one end, a sample loading pad, a mixing pad containing: 1) biotinylated RBD protein complexed with anti-biotin-conjugated to colloidal gold; 2) colloidal gold-conjugated chicken IgY as a control and reference standard. The nitrocellulose membrane strip has three stripes, the first of which is a recombinant chimeric protein consisting of the extracellular domain of human ACE2 fused to human-IgG1-Fc domain line (test line 1), a control/reference line (Ref Line) coated with anti-chicken IgY antibody and a total anti-RBD antibody line (test Line 2) coated with recombinant RBD protein.

The test specimen (15µl plasma) is dispensed into the sample well of the test cassette, and three drops of running buffer (phosphate buffered saline pH 7.4, 0.5% Tween20, 0.05% Sodium azide) are added to well B of the test cassette. For the test modified for whole blood, the test specimen (30µl) and running buffer are added to the same well. In both cases, the specimen mixes with the colloidal gold RBD conjugates upon sample addition and during migration by capillary action in the mixing pad, and then migrate along the nitrocellulose membrane. Anti-SARS-CoV-2 RBD antibodies, if present in the specimen, will bind to the RBD-biotin-Au conjugates. If no antibody is present, or if the antibodies bind to RBD protein but without capacity to interfere with ACE2 binding (*i*.*e*., lack of neutralization), then the maximum amount of RBD-gold will bind to the ACE2 line (test line 1). If neutralizing antibody is present in the specimen, less of the immunocomplex will bind to the ACE2 protein. In either case, as the sample continues to flow, the chicken IgY-gold will bind to the anti-chicken IgY line (control/reference line), and next, additional SARS-CoV-2 conjugate will flow to test line 2, where binding of antibody-antigen complexes to the same or related antigen on the test strip will indicate the presence of antibodies to SARS-CoV-2 that may or may not be neutralizing because ACE2 is not involved in this interaction. This indicates a SARS-CoV-2 RBD antibody positive test result.

The intensity of the control/reference line can be adjusted by varying the amount of anti-chicken IgY on the nitrocellulose and/or the amount of chicken IgY-gold at the time of manufacture, in order to reflect the best estimates for protective levels of antibody for immunity to SARS-CoV-2 infection. The RBD-biotin can be expressed using any variant sequence of RBD and is then titrated to determine an equivalent level of binding for the inhibition assay.

### Interpretation of test result

*(a) Valid assay* In addition to the presence of the reference line, if only test line 1 is developed and is more intense than the reference line, which is calibrated to reflect a defined level of intensity in test line 1, this result indicates levels of neutralising anti-RBD antibody in the sample are below the amount deemed to be neutralising based on the reference line. If test line 1 is less intense than the reference line, or very faint or invisible, this suggests that there is an adequate level of neutralising antibody present in the sample. Test line 2 may develop indicating the presence of anti-RBD antibody, indicating prior exposure to SARS-CoV-2 infection or RBD-containing vaccine, but this is not necessarily neutralising. These readings can be performed at a semi-quantitative level by eye as greater than or less than the reference line, as used in the visual interpretation of the Visitect® CD4 T-cell tests, or quantitatively with a wide dynamic range using an instrument such as the Axxin AX-2XS instrument. *(b) Invalid assay* If the C Line does not develop, the assay is invalid regardless of color development of the test line 1 and test line 2 as indicated below. In such cases, the assay should be repeated with a new device.

### Microneutralisation assay

Microneutralisation assays were performed as previously described ^18, 33^. SARS-CoV-2 isolate CoV/Australia/VIC01/2020^34^ was passaged in Vero cells and stored at −80C. Plasma was heat inactivated at 56C for 30 min, then serially diluted 1:20 to 1:10,240 before the addition of 100 TCID^50^ of SARS-CoV-2 in MEM/0.5% BSA and incubation at room temperature for 1 h. Residual virus infectivity in the plasma/virus mixtures was assessed in quadruplicate wells of Vero cells incubated in serum-free media containing 1 μg/ml of Tosyl phenylalanyl chloromethyl ketone (TPCK) trypsin at 37C and 5% CO^2^; viral cytopathic effect was read on day 5. The neutralizing antibody titre was calculated using the Reed–Muench method, as previously described^35^

### Statistics

Regression lines fitted in GraphPad Prism 9.0 using a semi-log non-linear fit, plotted with 95% CI bands. R^2^ goodness-of-fit displayed on graphs. For comparison of multiple groups, Kruskal-Wallis one-way ANOVA with Dunn’s multiple comparison test.

## Data Availability

All data referred to in the manuscript are available upon reasonable request

## Funding and Acknowledgements

We are grateful to the volunteers who provided samples for study. The authors would like to the Australian Red Cross Lifeblood for the supply of specimens from COVID-19 convalescent plasma donors. Australian Governments fund the Australian Red Cross Lifeblood to provide blood, blood products and services to the Australian community. The authors gratefully acknowledge the contribution to this work of the Victorian Operational Infrastructure Support Program received by the Burnet Institute. This work was supported by grants from the Department of Health and Human Services (DHHS) of the Victorian State Government; the Australian Research Council (ARC; CE140100011, CE140100036) the National Health and Medical Research Council of Australia (NHMRC; 1113293, 2002317 and 1116530), and Medical Research Future Fund (MRFF) Awards (2005544, 2002073, 2002132). THON and AKW were supported by an NHMRC Emerging Leadership Level 1 Investigator Grants (#1194036), HV was supported by an NHMRC APPRISE Research Fellowship (1116530), KK was supported by the NHMRC Leadership Investigator Grant (1173871), DLL was supported by a NHMRC Principal Research Fellowship (#1137285), KS and DAW are supported by NHMRC Investigator Grants (1177174 and 1174555). KS also received support from the A2 Milk Company and the Jack Ma Foundation. The Melbourne WHO Collaborating Centre for Reference and Research on Influenza is supported by the Australian Government Department of Health. DIG and SJK were supported by an NHMRC Senior Principal Research Fellowships (1117766).

## Author contributions

TSF, HV, NAG, SZ, MC, SR, RC, HD, SG, THON, FLM, LCR, DLD, ACC, PMH, ZM, KS, KK, BDW, DAW, HXT, AKW, JAJ, SJK performed experiments, provided samples or reagents and/or directed experimental work. DIG, DFJP, DAA conceived the study, and analysed the data. DIG and DAA co-wrote the paper and co-led the project.

## References

1. Huang, C. et al. 6-month consequences of COVID-19 in patients discharged from hospital: a cohort study. The Lancet 397, 220–232 (2021).

2. Poland, G.A., Ovsyannikova, I.G. & Kennedy, R.B. SARS-CoV-2 immunity: review and applications to phase 3 vaccine candidates. The Lancet 396, 1595–1606 (2020).

3. Forni, G., Mantovani, A. & Covid-19 Commission of Accademia Nazionale dei Lincei, R. COVID-19 vaccines: where we stand and challenges ahead. Cell Death Differ 28, 626–639 (2021).

4. Bond, K. et al. Evaluation of serological tests for SARS-CoV-2: Implications for serology testing in a low-prevalence setting. J Infect Dis (2020).

5. Wang, P. et al. Antibody Resistance of SARS-CoV-2 Variants B.1.351 and B.1.1.7. Nature (2021).

6. Khoury, D.S. et al. What level of neutralising antibody protects from COVID-19? Nature Medicine, in press (2021).

7. Khoury, D.S. et al. Measuring immunity to SARS-CoV-2 infection: comparing assays and animal models. Nat Rev Immunol 20, 727–738 (2020).

8. Piccoli, L. et al. Mapping neutralizing and immunodominant sites on the SARS-CoV-2 spike receptor-binding domain by structure-guided high-resolution serology. Cell (2020).

9. Wang, Q. et al. Structural and Functional Basis of SARS-CoV-2 Entry by Using Human ACE2. Cell 181, 894–904 e899 (2020).

10. Schmidt, F. et al. Measuring SARS-CoV-2 neutralizing antibody activity using pseudotyped and chimeric viruses. J Exp Med 217 (2020).

11. Tan, C.W. et al. A SARS-CoV-2 surrogate virus neutralization test based on antibody-mediated blockage of ACE2-spike protein-protein interaction. Nat Biotechnol 38, 1073–1078 (2020).

12. Ju, B. et al. Human neutralizing antibodies elicited by SARS-CoV-2 infection. Nature 584, 115–119 (2020).

13. Abe, K.T. et al. A simple protein-based surrogate neutralization assay for SARS-CoV-2. JCI Insight 5 (2020).

14. Byrnes, J.R. et al. A SARS-CoV-2 serological assay to determine the presence of blocking antibodies that compete for human ACE2 binding. medRxiv (2020).

15. Walker, S.N. et al. SARS-CoV-2 Assays To Detect Functional Antibody Responses That Block ACE2 Recognition in Vaccinated Animals and Infected Patients. J Clin Microbiol 58 (2020).

16. Lake, D.F. et al. Development of a Rapid Point-Of-Care Test that Measures Neutralizing Antibodies to SARS-CoV-2. medRxiv (2021).

17. Tian, X. et al. Potent binding of 2019 novel coronavirus spike protein by a SARS coronavirus-specific human monoclonal antibody. Emerg Microbes Infect 9, 382–385 (2020).

18. Koutsakos, M. et al. Integrated immune dynamics define correlates of COVID-19 severity and antibody responses. 2, 100208 (2021).

19. Muhi, S. et al. Multi-site assessment of rapid, point-of-care antigen testing for the diagnosis of SARS-CoV-2 infection in a low-prevalence setting: A validation and implementation study. The Lancet Regional Health - Western Pacific 9 (2021).

20. Luchters, S. et al. Field Performance and Diagnostic Accuracy of a Low-Cost Instrument-Free Point-of-Care CD4 Test (Visitect CD4) Performed by Different Health Worker Cadres among Pregnant Women. J Clin Microbiol 57 (2019).

21. Seow, J. et al. Longitudinal observation and decline of neutralizing antibody responses in the three months following SARS-CoV-2 infection in humans. Nat Microbiol 5, 1598–1607 (2020).

22. Long, Q.X. et al. Clinical and immunological assessment of asymptomatic SARS-CoV-2 infections. Nat Med 26, 1200–1204 (2020).

23. Marot, S. et al. Rapid decline of neutralizing antibodies against SARS-CoV-2 among infected healthcare workers. Nat Commun 12, 844 (2021).

24. Tan, H.X. et al. Immunogenicity of prime-boost protein subunit vaccine strategies against SARS-CoV-2 in mice and macaques. Nat Commun 12, 1403 (2021).

25. Greaney, A.J. et al. Comprehensive mapping of mutations to the SARS-CoV-2 receptor-binding domain that affect recognition by polyclonal human serum antibodies. bioRxiv (2021).

26. Thomson, E.C. et al. Circulating SARS-CoV-2 spike N439K variants maintain fitness while evading antibody-mediated immunity. Cell 184, 1171–1187 e1120 (2021).

27. Pham, M.D. et al. Performance of a Novel Low-Cost, Instrument-Free Plasma Separation Device for HIV Viral Load Quantification and Determination of Treatment Failure in People Living with HIV in Malaysia: a Diagnostic Accuracy Study. Journal of clinical microbiology 57, e01683–01618 (2019).

28. Buchholz, U.J. et al. Contributions of the structural proteins of severe acute respiratory syndrome coronavirus to protective immunity. Proc Natl Acad Sci U S A 101, 9804–9809 (2004).

29. McMahan, K. et al. Correlates of protection against SARS-CoV-2 in rhesus macaques. Nature 590, 630–634 (2021).

30. Robbiani, D.F. et al. Convergent antibody responses to SARS-CoV-2 in convalescent individuals. Nature 584, 437–442 (2020).

31. Lau, E.H.Y. et al. Neutralizing antibody titres in SARS-CoV-2 infections. Nat Commun 12, 63 (2021).

32. Vazquez-Lombardi, R. et al. Transient expression of human antibodies in mammalian cells. Nat Protoc 13, 99–117 (2018).

33. Juno, J.A. et al. Humoral and circulating follicular helper T cell responses in recovered patients with COVID-19. Nat Med (2020).

34. Caly, L. et al. Isolation and rapid sharing of the 2019 novel coronavirus (SARS-CoV-2) from the first patient diagnosed with COVID-19 in Australia. Med J Aust 212, 459–462 (2020).

35. Subbarao, K. et al. Prior infection and passive transfer of neutralizing antibody prevent replication of severe acute respiratory syndrome coronavirus in the respiratory tract of mice. J Virol 78, 3572–3577 (2004).

